# Higher pharyngeal epithelial gene expression of Angiotensin-Converting Enzyme-2 in upper respiratory infection patients

**DOI:** 10.1101/2020.06.27.20141754

**Authors:** Mingjiao Zhang, Lingyan Du, Oluwasijibomi Damola Faleti, Jing Huang, Gang Xiao, Xiaoming Lyu

## Abstract

We analyzed the expression of ACE2 in pharyngeal epithelium and examined its relationship with clinical features and serological parameters in the upper respiratory infection (URI) patients. The expression of ACE2 were significantly higher in URI patients than in healthy controls individuals, and positively correlated with age and body temperature.

---

Co-infection with severe acute respiratory syndrome coronavirus 2 (SARS-CoV-2) and different respiratory pathogens has been reported recently[1]. It is hypothesized that the higher risk among coronavirus disease 2019 (COVID-19) patients with co-infection is due to differential expression of angiotensin-converting enzyme 2 (ACE2), the receptor that SARS-CoV-2 uses for host entry. We analyzed ACE2 gene expression in the pharyngeal epithelium of upper respiratory infection (URI) patients and healthy individuals.

## METHORDS

we conducted a retrospective examination of pharyngeal epithelium from individuals encountered within the Third Affiliated Hospital, Southern Medical University, China, from March 2020 to May 2020. Samples were collected from125 cases of URI patients and 52 age- and sex-matched healthy individuals with no infection for research on receptor of SARS-CoV-2, all participants have negative tests for SARS-CoV-2. Written informed consent was obtained from participants. Pharyngeal epithelium of throat swabs was collected and immediately placed in RNA stabilization fluid. Total RNA was extracted from pharyngeal epithelium and stored at −80 °C. Real-time transcription-polymerase chain reaction analysis (RT-PCR) was used to determine expression of ACE2 gene. The relative expression level of ACE2 was normalized to the internal control GAPDH expression and calculated by the comparative C_T_ (△△C_T_) method. All data were statistically analyzed using Graph Pad Prism5 (version 5.0) software. Quantitative data are expressed as the mean±SD. Data with a Gaussian distribution was analyzed using an unpaired t test or one-way analysis of variance (ANOVA). Spearman’s correlation test was performed to assess the correlation of ACE2 gene and clinical variables. *P* values less than 0.05 were considered statistically significant.

Considering the evidence that ACE2 is the main host cell receptor of SARS-CoV-2 and plays a crucial role in the entry of virus into the cell to cause the final infection, ACE2 gene expression was the focus of this study.

## RESULTS

The cohort of 177 individuals aged 1 to 95 years, the median age of all URI patients was 33.69±15.88years, 57.6% were males (72/125). Among all URI patients, 65.6%(82/125) got a fever with body temperature over 37.4°C. The expression level of ACE2 gene was significantly higher in URI patients (n=125) than in Healthy control (HC) individuals (n=52) (*p*<0.0001, **Fig.1**). ACE2 gene expression level was significantly and positively correlated with age (r=0.1799, *p*=0.0447) and body temperature (r=0.1927, *p*=0.0427) (**Table 1**).

**Figure 1.**
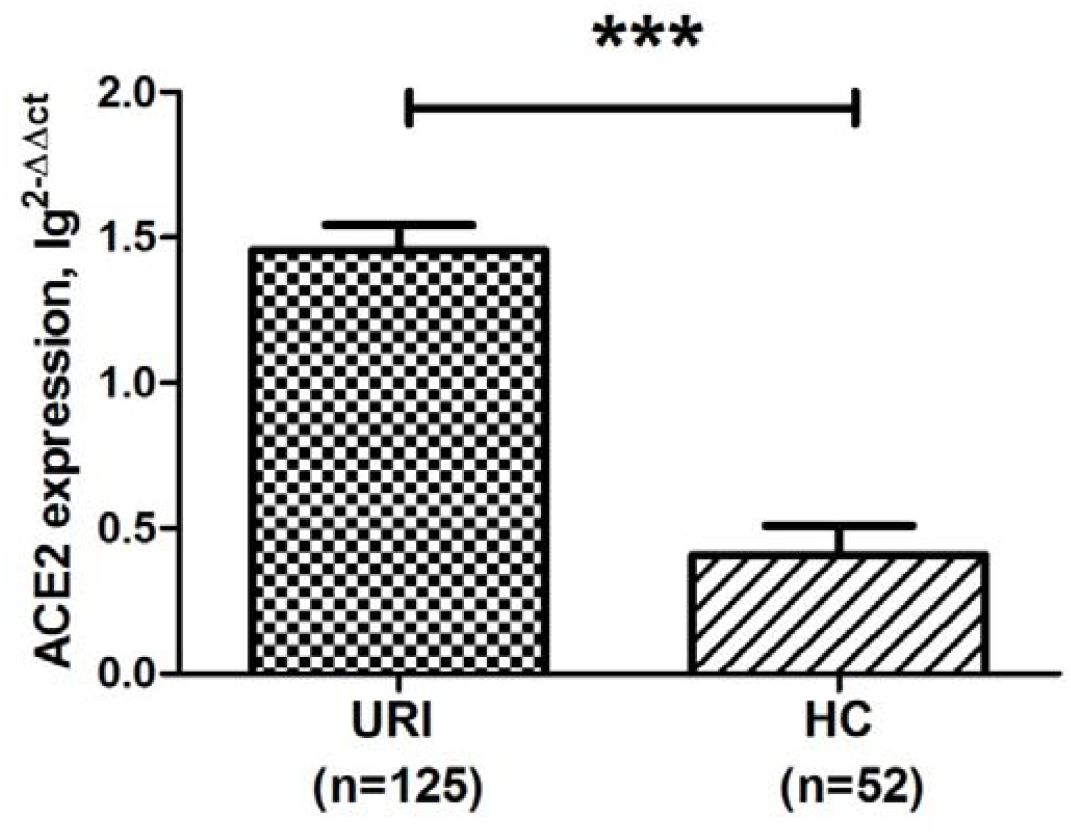
The increased levels of ACE2 gene in pharyngeal epithelium of URI patients.

**Table 1.**
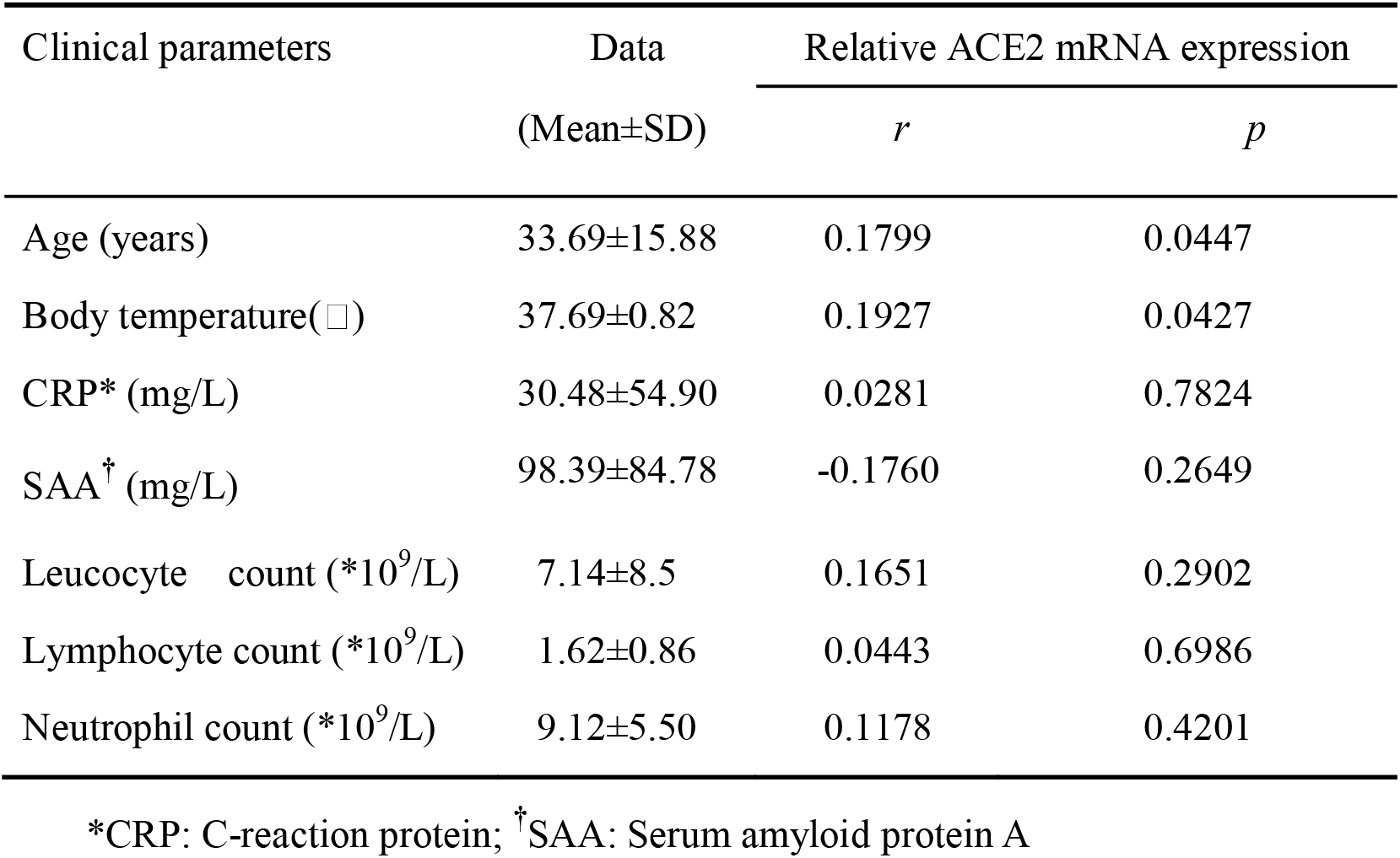
Association of ACE2 gene with clinical pathological parameters of URI patients

| Clinical parameters | Data<br>(Mean±SD) | Relative ACE2 mRNA expression |  |
| --- | --- | --- | --- |
|  |  | *r* | *p* |
| Age (years) | 33.69±15.88 | 0.1799 | 0.0447 |
| Body temperature(□) | 37.69±0.82 | 0.1927 | 0.0427 |
| CRP* (mg/L) | 30.48±54.90 | 0.0281 | 0.7824 |
| SAA<sup>†</sup> (mg/L) | 98.39±84.78 | -0.1760 | 0.2649 |
| Leucocyte count (*10<sup>9</sup>/L) | 7.14±8.5 | 0.1651 | 0.2902 |
| Lymphocyte count (*10<sup>9</sup>/L) | 1.62±0.86 | 0.0443 | 0.6986 |
| Neutrophil count (*10<sup>9</sup>/L) | 9.12±5.50 | 0.1178 | 0.4201 |

\*CRP: C-reaction protein; <sup>†</sup>SAA: Serum amyloid protein A

Data are means and 95% confidence intervals (error bars) for ACE2 gene expression in the pharyngeal epithelium of URI patients and HC individuals, **\*\*\*** (*p*<0.0001). Data were log-transformed by taking the base 10 logarithm to account for the skewness. After the above processing, the treated data were normal distributed and had constant variances.

## DISCUSSION

In this study, we showed that ACE2 gene level was significantly upregulated in the pharyngeal epithelium of URI patients. We also found that ACE2 gene level was significantly and positively correlated with age and body temperature. So far as we know, there is few reports demonstrating that ACE2 gene level in the pharyngeal epithelium is increased in URI patients. A recent study reported ACE2 is an interferon-stimulated gene in human airway epithelial cells, which may help explain increasing coinfections with SARS-CoV-2 and other respiratory pathogens(2).

The Chinese experts from China-Japan Friendship Hospital have reported a case coinfection with influenza A virus and SARS-CoV-2[3]. A recent study also showed that a total of 5 of 115 patients confirmed with COVID-19 was diagnosed with influenza virus infection[4]. There are still other case reports have described coinfection with other respiratory pathogens[1]. Those raise the concerns that there might be mixed infections of seasonal influenza and the novel coronavirus. Measures should be taken to enhance the respiratory infectious diseases surveillance systems and avoid lethal secondary infections.

This study provides novel results on ACE2 gene expression in the pharyngeal epithelium and its relationship with URI disease. The molecular mechanism that how to activate ACE2 in URI patients might need to be future explored.

## Data Availability

Not applicable.

## Acknowledgments

*Miss*. Mingjiao Zhang and *Miss*. Lingyan Du contribute equally to this work. *Miss*. Mingjiao Zhang is a *Ph*.*D*. candidate of Southern Medical University. While completing this work, they are engaging in nucleic acid screening of SARS-CoV-2. Their primary research interests are infectious diseases. We thank our colleagues from the department of laboratory medicine, The Third Affiliated Hospital, Southern Medical University for their kindly help.

## Disclaimer

The findings and conclusions in this report are those of the authors and do not necessarily represent the official position of the Centers for Disease Control and Prevention.

## References

1. Khaddour K, Sikora A, Tahir N, Nepomuceno D, Huang T. Case Report: The Importance of Novel Coronavirus Disease (COVID-19) and Coinfection with Other Respiratory Pathogens in the Current Pandemic. AM J TROP MED HYG 2020 2020-06-01;102(6):1208–9.

2. Ziegler C, Allon SJ, Nyquist SK, et al. SARS-CoV-2 Receptor ACE2 Is an Interferon-Stimulated Gene in Human Airway Epithelial Cells and Is Detected in Specific Cell Subsets across Tissues. CELL 2020 2020-05-28;181(5):1016–35.

3. Wu X, Cai Y, Huang X, et al. Co-infection with SARS-CoV-2 and Influenza A Virus in Patient with Pneumonia, China. EMERG INFECT DIS 2020 2020-06-01;26(6):1324–6.

4. Ding Q, Lu P, Fan Y, Xia Y, Liu M. The clinical characteristics of pneumonia patients coinfected with 2019 novel coronavirus and influenza virus in Wuhan, China. J MED VIROL 2020 2020-03-20.

